# Effects of a preconception lifestyle intervention on cardiorespiratory fitness and glycaemic outcomes in people with increased risk of gestational diabetes: Secondary findings from the BEFORE THE BEGINNING randomised controlled trial

**DOI:** 10.1101/2025.11.25.25340951

**Authors:** MAJ Sujan, HMS Skarstad, G Rosvold, HN Lund, AP Oladewa, SL Fougner, T Follestad, KÅ Salvesen, T Moholdt

**Author notes:** Correspondence to: Moholdt T, Postal address: NTNU, Faculty of Medicine and Health Sciences, Department of Circulation and Medical Imaging, Post box 8905, 7491 Trondheim, Norway,Twitter (X) handle: @trinemoholdt. E-mail addresses: MAJS, HMSS, GR, HNL, APO, SLF, TF, KÅS, TM. Trial registration: ClinicalTrials.gov NCT04585581.

## Abstract

**Objectives:** Obesity rates are rising globally, and many females of reproductive age are physically inactive, which negatively affect future maternal and neonatal health. In this secondary analysis, we determined the effect of a combined preconception lifestyle intervention on cardiorespiratory fitness and glycaemic outcomes after 7 weeks.

**Methods:** In the BEFORE THE BEGINNING randomised controlled trial, we randomised females who were at risk of gestational diabetes mellitus and planning pregnancy 1:1 to intervention or control. The intervention consisted of time-restricted eating (TRE) and exercise training and spanned from inclusion preconception throughout pregnancy. TRE involved restricting energy intake to ≤ 10-h/day on ≥ 5 days per week. Exercise volume was based on a heart-rate-based metric (Personal Activity Intelligence, PAI), with the goal of ≥ 100 weekly PAI-points. The main outcomes of interest in this secondary analysis were cardiorespiratory fitness and glycaemic outcomes after 7 weeks of intervention in the preconception period.

**Results:** Among 167 randomised participants, we included 166 in the intention-to-treat analysis. After 7 weeks of intervention, there was no between-group difference in relative VO_2_peak (0.8 mLkg^-1^min^-1^; 95% CI -0.2 to 1.9, *P* = 0.10) or glucose area under the curve (0.0 mmol/L; 95% confidence interval, CI -0.1 to 0.2, *P* = 0.76).

**Conclusion:** We found no evidence of effects of 7 weeks of combined TRE and exercise training on cardiorespiratory fitness or glycaemic outcomes in people with increased risk of gestational diabetes.

## INTRODUCTION

Global rates of physical inactivity continue to rise in parallel with increasing prevalence of obesity and overweight [1,2]. Females of reproductive age are no exception; most remain physically inactive before and during pregnancy [2,3]. Evidence consistently shows that a healthy diet and regular physical activity protect maternal health both before and during pregnancy[4,5]. However, antenatal lifestyle interventions have produced only limited effects on maternal and neonatal outcomes [6,7]. Consequently, researchers have called for lifestyle interventions to begin prior to conception and emerging evidence supports this approach [8,9]. Despite these calls, few trials have evaluated preconception lifestyle interventions, and the literature remains divided on which strategies most effectively improve cardiometabolic health before pregnancy [10–12].

Time-restricted eating (TRE) is a form of intermittent fasting that restricts daily energy intake to a certain time-window. TRE improves several cardiometabolic outcomes in people who are overweight or have obesity [13]. High-intensity interval training (HIIT), which alternates short bouts of vigorous exercise with brief recovery periods, increases peak oxygen uptake (VO_2_peak) more effectively than continuous moderate-intensity training [14] and also improves cardiometabolic outcomes in this population [15,16]. Combining TRE with HIIT produces greater cardiometabolic improvements than either intervention alone or no intervention [15]. These time-efficient and practical strategies are particularly suitable for people of reproductive age who face unique barriers to maintain healthy lifestyle behaviours [17].

The BEFORE THE BEGINNING randomised controlled trial (RCT) evaluated a preconception lifestyle intervention combining ≤ 10-h TRE and exercise training in people at increased risk of gestational diabetes mellitus (GDM) [18]. In this report, we present results from the first 8 weeks of the trial, including cardiorespiratory fitness, continuous glucose monitoring (CGM), physical activity, and participant-reported hunger and appetite. We hypothesised that 7 weeks of combined TRE and exercise would improve cardiorespiratory fitness and glycaemic outcomes compared with a no-intervention control group.

## METHODS

### Trial design and participants

The BEFORE THE BEGINNING trial was a two-arm, parallel-group RCT conducted at the Norwegian University of Science and Technology (NTNU), Trondheim, Norway. Recruitment took place from October 2020 to May 2023, with participants randomised 1:1 to either the intervention or a no-intervention control group. The trial was registered in ClinicalTrials.gov (NCT04585581) on September 25, 2020. We have previously published the study protocol [18] and main trial results [19].

Eligible participants in the BEFORE THE BEGINNING trial were females aged 18-39 years who planned to conceive within 6-12 months and understood oral and written Norwegian or English. Participants were required to have at least one risk factor for GDM [20]: Body mass index (BMI) ≥ 25 kg/m^2^, previous GDM, first-degree relative with diabetes, prior delivery of a newborn ≥ 4.5 kg, or non-European ethnicity (defined as having one or both parents originating outside Europe). Exclusion criteria included ongoing pregnancy, attempts to conceive for ≥ six cycles before enrolment, diagnosed diabetes mellitus or cardiovascular disease, shift work involving > 2 nights/week, high intensity exercise > 2 sessions/week in the previous 3 months, a habitual eating window of ≤ 12 h/day, or any other condition deemed incompatible with participation by the investigators. We recruited participants through social media, public advertising, hospital and university websites. Starting in November 2022, we additionally recruited participants via electronic invitations distributed using population data from the Norwegian Tax Administration. Full details of eligibility criteria and recruitment procedures are provided in the published protocol [18].

### Randomisation and blinding

Following baseline assessments, we randomised the participants 1:1 to either an intervention or a standard care control group, stratified by previous GDM diagnosis. A technician at The Clinical Research Unit (Klinforsk) at NTNU and St. Olav’s Hospital (Trondheim, Norway) generated the randomisation sequence using a computer-based random number generator (WebCRF3). We used variable block sizes for randomisation and concealed the group allocation until interventions were assigned. The investigators enrolled and assigned the participants to groups. The participants, study personnel or outcome assessors were not blinded to the group allocation.

### Intervention

Participants in the intervention group received a combined TRE and exercise training intervention. We instructed them to restrict daily energy intake to a ≤ 10-h window on ≥ 5 days/week, with the last energy intake no later than 19:00 h. On the remaining 2 days, participants were free to choose their eating window. We did not give any additional dietary counselling, and participants could consume calorie-free beverages outside the eating window. The exercise intervention consisted of endurance training, guided by the heart rate(HR)-based Personal Activity Intelligence (PAI) algorithm [21]. We instructed the participants to maintain ≥ 100 PAI-points per rolling week, a threshold associated with reduced risk of cardiovascular disease [21,22]. Because higher HR yield higher PAI scores, this approach encouraged high-intensity exercise. The exercise training was mainly unsupervised, but we offered additional supervised sessions on treadmill or stationary bikes at the study facility at first and 3rd intervention week and on request. We collected PAI-data throughout the intervention through smartwatches worn by the participants in the intervention group, and we reinforced adherence via text message reminders.

We asked the participants in the control group to maintain their habitual diet and physical activity. All study participants received a brochure from the national health authorities outlining physical activity and dietary recommendations for the preconception and pregnancy periods.

### Outcomes

We conducted assessments twice during the preconception period: at baseline and intervention week 7 (Figure 1). The outcomes presented in this report are described below. In the first week following initial laboratory assessments and randomisation, participants in both groups continued with their habitual lifestyle (Figure 1).

**Figure 1.**
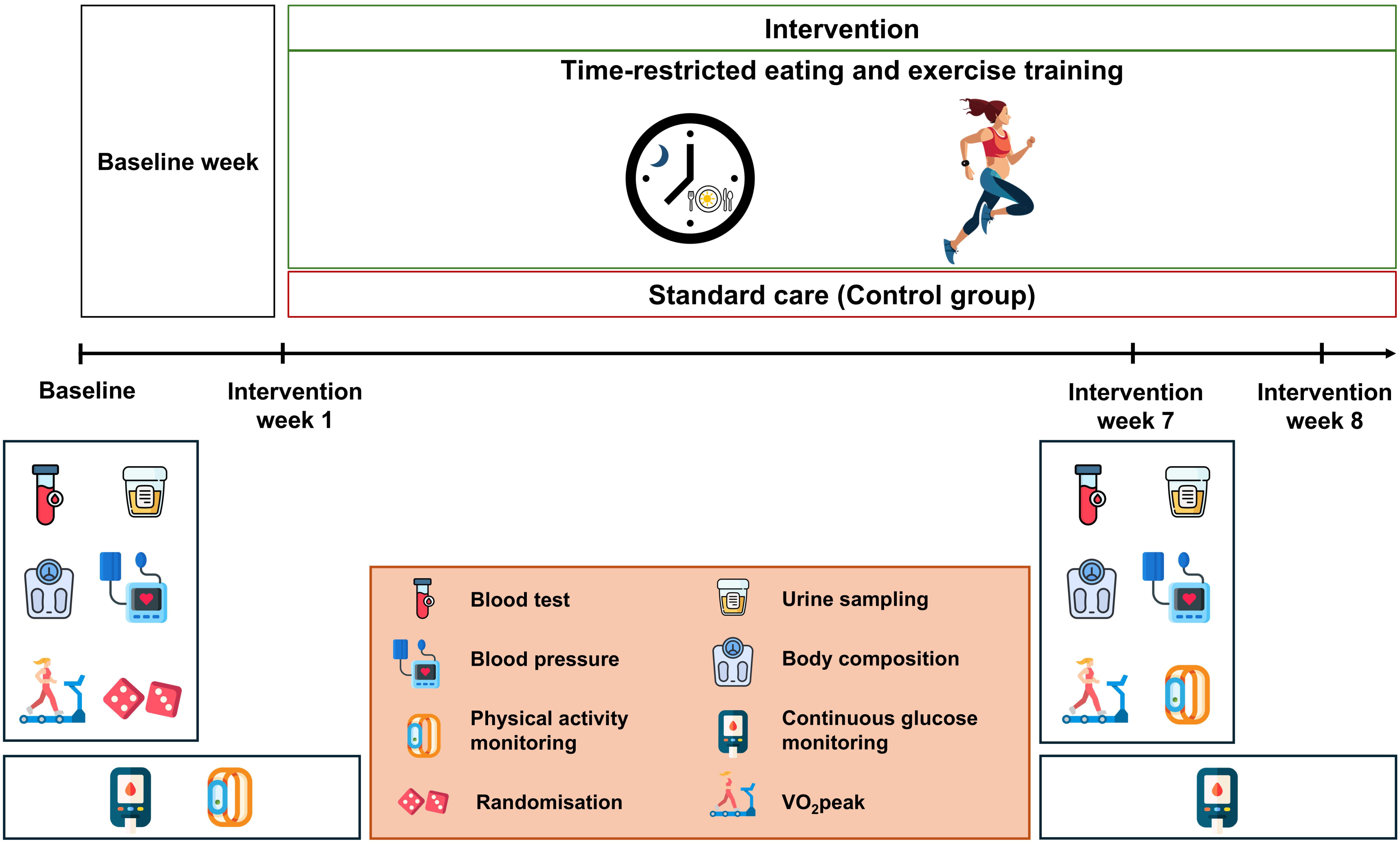
Assessments in the preconception period. Following baseline assessments, we randomised the participants 1:1 into the intervention or a control group. Additionally, we collected continuous glucose monitoring (CGM) and physical activity data from all participants during the no-intervention baseline week and first intervention week. We conducted the assessments again at intervention week 7 and CGM measurements were repeated for 14 days. The participants recorded their daily window of energy intake and subjective rating of hunger and appetite for 4 days (three weekdays and one weekend day) in the study handbook at baseline week and intervention week 7.

#### Cardiorespiratory fitness

The participants completed individualised, graded treadmill tests to measure VO_2_peak using indirect calorimetry (Metalyzer II, Cortex, Germany) at baseline. The test was repeated in no-pregnant participants at intervention week 7. Each test began with a 10-min warm-up at moderate intensity, and at a rating of 11-13 on the Borg (6-20) rating of perceived exertion [23]. We increased the speed (0,5-1,0 km/h) or incline (1-2%) every 1-2 mins, and participants exercised until they reached volitional exhaustion. Since our participants were previously untrained, we did not expect them to reach a plateau in O_2_ uptake and/or respiratory exchange ratio (RER) of ≥ 1,10, which is used to estimate maximum oxygen uptake (VO_2_max) [24]. Therefore, we report VO_2_peak, defined as the average of three highest consecutive 10-s O_2_ uptake measurements during the test, both as absolute (Lmin^-1^) and relative (mLkg^-1^min^-1^) VO_2_peak values. We estimated HR_max_ as the highest HR value recorded by chest strap HR monitors during the test [24]. We then assessed HR recovery as the reduction in HR from HR_max_ to the value recorded 1 min after the test ended [25].

#### Continuous glucose monitoring

The participants wore a CGM sensor (FreeStyle Libre 1, Abbott Diabetes Care, Norway) for 14 days starting from both the visits at baseline and intervention week 7. The first period comprised of 7 days of habitual diet-exercise behaviour, followed by the first 7 days of the intervention period. We instructed the participants to scan the sensors with CGM readers at least four times/day for calibration. To minimise bias, we covered the monitor screens. We processed the raw CGM data using Microsoft Excel and used Glyculator 3.0 [26] to estimate 24 h, daytime (06:00-00:00) and nocturnal (00:01-05:59) mean glucose, glucose area-under-the-curve (AUC), coefficient of variation (CV), and mean amplitude of glucose excursions (MAGE).

#### Physical activity monitoring

Intervention participants wore smartwatches (Amazfit GTS, Huami, China/Polar Ignite 2, Polar, Finland) throughout the study period, which converted HR data into PAI scores and uploaded to a secure online portal accessible by the investigators. All participants wore activity armbands (BodyMedia SenseWear, Pennsylvania, USA) for 14 days at baseline, covering the baseline week of habitual lifestyle and the first intervention week. We processed data from activity armbands and smartwatches in Microsoft Excel. In this report, we present daily physical activity levels as metabolic equivalents of tasks (METs), energy expenditure, time spent being sedentary and in light, moderate, vigorous, and very vigorous activity, total physical activity time, and daily step count. Exercise adherence was evaluated using weekly PAI-points, as reported previously [19].

#### Self-reported time-window of energy intake and hunger and appetite

After randomisation, we provided the participants a study handbook where they completed a 4-day registration (three weekdays and one weekend day) at baseline and intervention week 7. They recorded the timing of first and last energy intake and subjective feeling of morning and evening hunger and appetite on 10 cm visual analogue scales. These outcomes are detailed in the published trial protocol [18].

### Sample size

The sample size calculation was based on the primary outcome measure: 2-h plasma glucose during a 75 g oral glucose tolerance test (OGTT) in gestational week 28 [18]. Power analysis showed that 74 participants (37 in each group) needed to reach gestational week 28 to achieve a statistical power of 0.90 at a 0.05 significance level. Recruitment was stopped after 167 participants were enrolled, at which point 47 participants in each group had reached gestational week 12. Full details of the sample size calculation are provided in the trial protocol [18].

### Statistical methods

We used linear mixed models to estimate differences between groups, with results presented as estimated effects for the intervention group relative to the control group, accompanied by 95% confidence intervals (CI) and *P*-values. Normality assumptions were evaluated by visual inspection of QQ-plots. Given the exploratory nature of the secondary outcomes, the significance threshold was pragmatically set to 0.01 to reduce the risk of type I errors due to multiple comparisons. We modified the prespecified per-protocol analysis plan to reflect adherence during the first 7 weeks of preconception intervention[18]. Participants were included if they achieved ≥ 75 weekly PAI points and reported a ≤ 10-hour eating window on at least 2 of 4 days during this period. We used IBM SPSS Statistics v.29.0.1 (IBM Corp, Armonk, NY, USA) for statistical analyses.

### Modifications to the trial protocol

We made several protocol modifications after trial commencement in October 2020. In June 2021, we introduced supervised exercise sessions to all intervention participants shortly after enrolling. From November 2022, we expanded screening using population data from the Norwegian Tax administration to improve outreach. During this period, we modified our exclusion criteria by adding “inability to undergo one or both interventions” and removing “planned assisted fertilisation”. In June 2023, we updated the study devices and applications: changed smartwatches from Amazfit GTS to Polar Ignite 2 and replaced the Zepp and Memento applications to Polar Flow and Mia Health to enhance PAI data capture. Additional modifications are detailed in the protocol article [18].

### Patient and public involvement

In our study, we involved users (reproductive-aged women with overweight/obesity) during the planning phase and implementation, but we did not involve the participants in the recruitment process. We conducted interactive online meetings and workshops to explore barriers to participation, enhance motivation and engagement, and discuss data collection, recruitment strategies, and adherence during the study.

## RESULTS

We enrolled a total of 167 participants in the BEFORE THE BEGINNING trial (intervention, *n* = 84, control, *n* = 83). The first participant was enrolled on October 2, 2020, and the last participant on May 12, 2023. We excluded one participant in the intervention group after randomisation due to detection of prepregnancy diabetes, leaving 166 participants in the intention-to-treat analysis. Baseline characteristics were comparable between groups (Table 1). Participant flow during the preconception period is shown in Figure 2. These characteristics are also reported in detail in the primary article [19].

**Figure 2:**
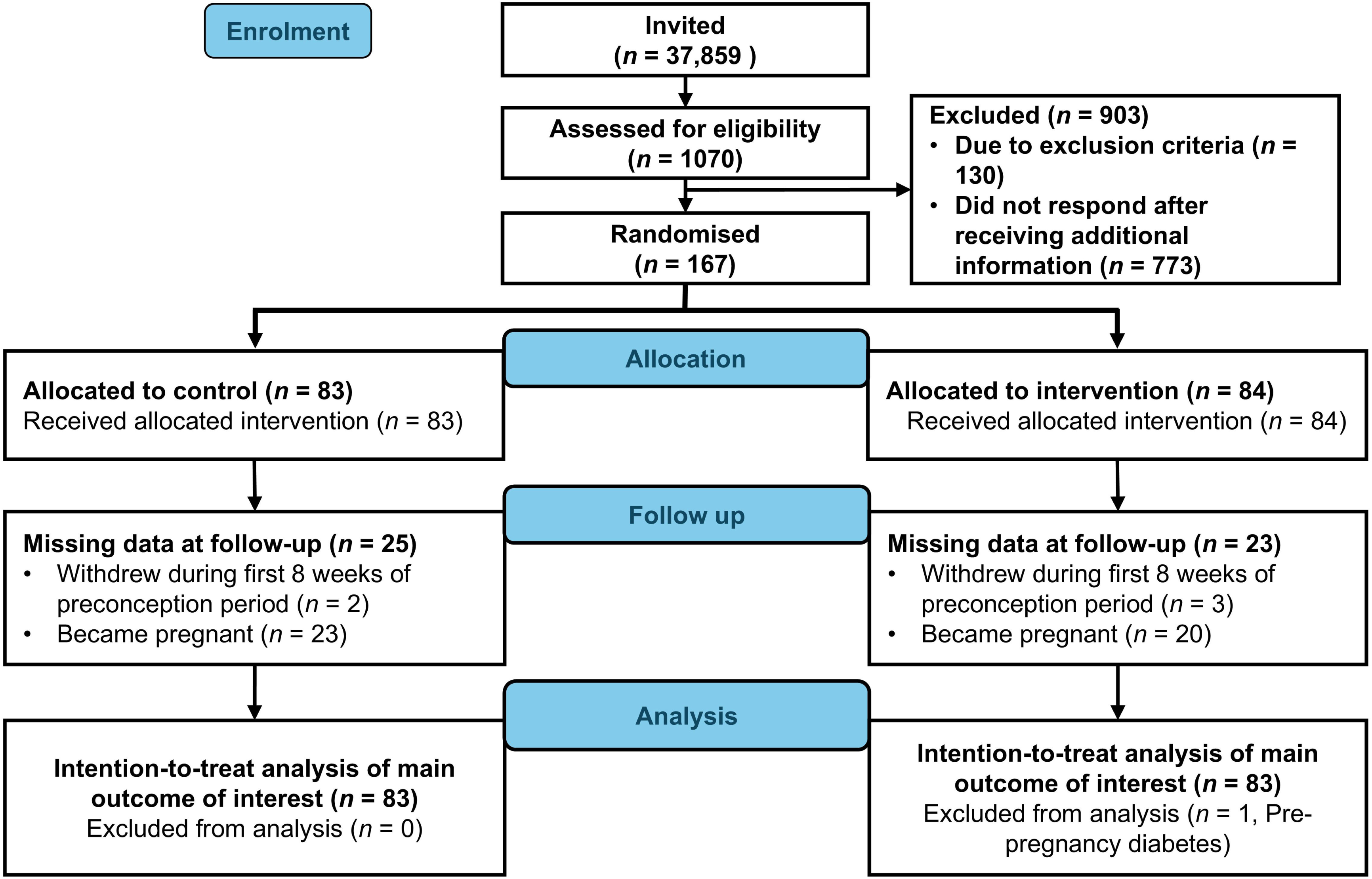
Participant flow in the trial (CONSORT Flow diagram).

**Table 1.** Baseline characteristics of participants, according to group allocation.

| Characteristics | Control (*n* = 83) | Intervention (*n* = 83) |
| --- | --- | --- |
| Age, years | 30.3 | 30.2 |
| Height, cm | 167.3 (5.6) | 166.4 (7.2) |
| Weight, kg | 81.5 (13.3) | 81.1 (16.0) |
| 24-h mean glucose, mmol/L | 4.9 (0.6) | 4.8 (0.5) |
| 24-h glucose AUC, mmol·h/L | 4.5 (0.5) | 4.4 (0.5) |
| 24-h CV, % | 16.9 (3.8) | 16.9 (3.8) |
| 24-h MAGE, mmol/L | 1.6 (0.4) | 1.6 (0.4) |
| Absolute VO<sub>2</sub>peak, Lmin<sup>-1</sup> | 2.9 (0.4) | 3.0 (0.5) |
| Relative VO<sub>2</sub>peak, mLkg<sup>-1</sup>min<sup>-1</sup> | 35.9 (5.6) | 37.1 (6.6) |
| Maximum rate of perceived exertion | 19.2 (0.7) | 19.3 (0.7) |
| Maximum respiratory exchange ratio | 1.1 (0.1) | 1.1 (0.1) |
| Heart rate recovery, beats per min | 31.2 (11.8) | 30.1 (9.0) |
| PAI-points |  | 75.9 (61.8) |
| Parity, *n* (%) |  |  |
| 0 | 45 (55) | 47 (59) |
| 1 | 30 (37) | 29 (36) |
| 2 | 7 (9) | 4 (5) |

Data are presented as means of observed values with standard deviations (SDs) or frequencies (*n*) and percentages (%) of participants. Missing: number of participants with missing data in each group is 0 to 3 for all variables. AUC = Area under the curve, CV = Coefficient of variation, and MAGE = Mean amplitude of glucose excursions, PAI = Personal activity intelligence points. PAI-points were only recorded in the intervention group.

### Cardiorespiratory fitness and physical activity monitoring

After 7 weeks of intervention, there were no statistically significant between-group differences in absolute (0.1 Lmin^-1^; 95% CI 0.0 to 0.1, *P =* 0.17) or relative VO_2_peak (0.8 mLkg^-1^min^-1^; 95% CI -0.2 to 1.9, *P =* 0.10) (Figure 3). No significant differences were observed in rate of perceived exertion or HR recovery (Supplementary Table 1). Data from activity armband during the first 2 weeks showed that the intervention participants accumulated more daily minutes of very vigorous activity in week 1 (0.8 min; 95% CI -0.3 to 1.4, *P =* 0.001) compared with the controls (Table 3). The remaining physical activity outcomes did not differ significantly between-groups.

**Figure 3.**
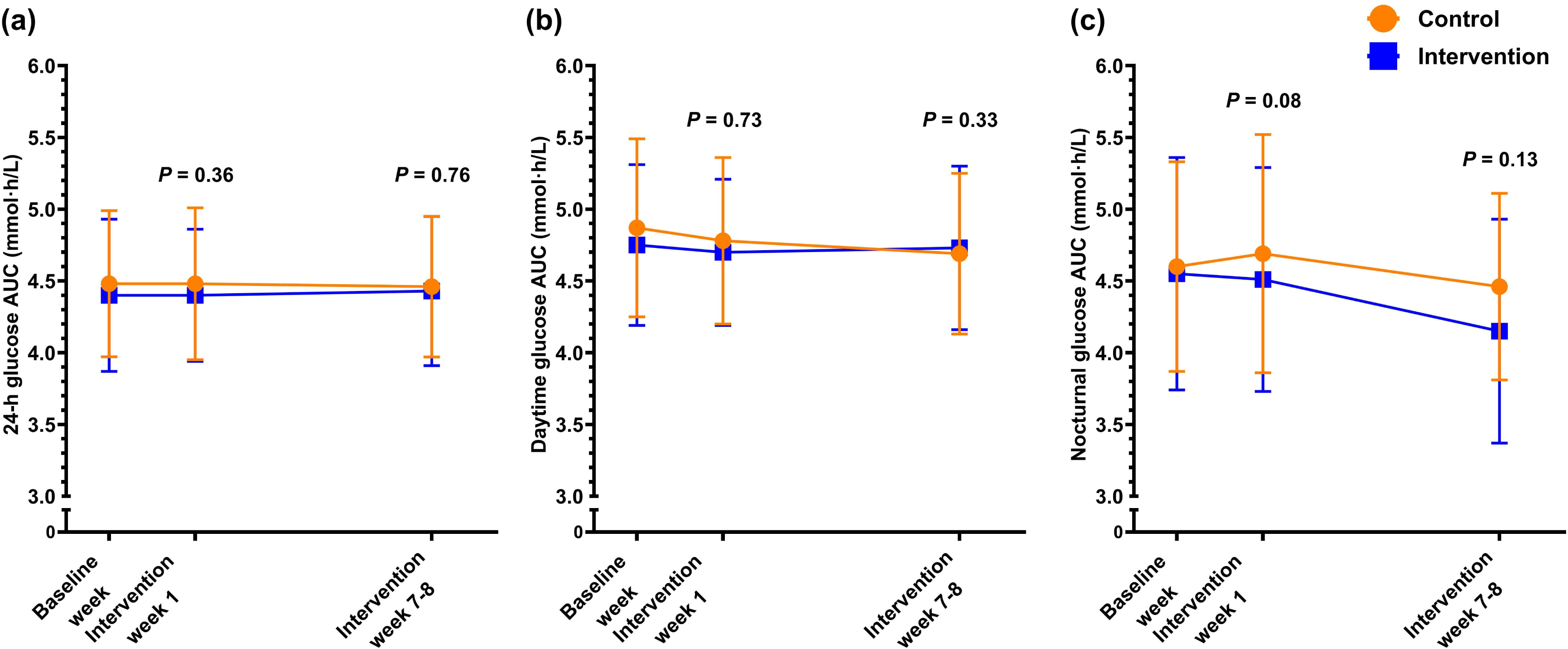
VO_2_peak measured at baseline and intervention week 7 according to group. Data are presented as observed means for intention-to-treat population, error bars show mean ± 1 standard deviation (SD), and the lines represent changes in each group from baseline. The *P*-values were calculated for a test of between group differences, using a linear mixed model.

**Table 3.** Daily physical activity outcomes estimated using activity armbands.

| Outcome | Time | Control | Intervention | Difference<br>(intervention- control) |  |
| --- | --- | --- | --- | --- | --- |
|  |  | Mean (SD) | Mean (SD) | Est. effect (95% CI) | P |
| Physical activity level, METs | Baseline week | 1.3 (0.2) | 1.3 (0.2) |  |  |
|  | Int. week 1 | 1.2 (0.2) | 1.3 (0.2) | 0.0 (0.0 to 0.1) | 0.39 |
| Daily energy expenditure, kJ | Baseline week | 9347.5 (1332.6) | 9207.7 (1461.9) |  |  |
|  | Int. week 1 | 9369.2 (1293.7) | 9458.3 (1234.0) | 87.4 (-199.1 to 374.0) | 0.55 |
| Sedentary time, min | Baseline week | 1068.1 (157.7) | 1090.0 (168.0) |  |  |
|  | Int. week 1 | 1115.4 (174.8) | 1097.2 (180.4) | -16.6 (-60.4 to 27.3) | 0.46 |
| Light intensity activity, min | Baseline week | 153.6 (76.7) | 151.9 (84.5) |  |  |
|  | Int. week 1 | 144.8 (77.4) | 149.2 (79.7) | 5.9 (-11.8 to 23.5) | 0.51 |
| Moderate intensity activity, min | Baseline week | 48.2 (29.4) | 55.3 (40.2) |  |  |
|  | Int. week 1 | 42.3 (29.1) | 56.2 (28.9) | 6.8 (-2.0 to 15.6) | 0.13 |
| Vigorous intensity activity, min | Baseline week | 3.5 (6.6) | 6.2 (9.8) |  |  |
|  | Int. week 1 | 3.5 (6.7) | 6.1 (8.6) | 0.3 (-2.0 to 2.7) | 0.79 |
| Very vigorous intensity activity, min | Baseline week | 0.3 (0.8) | 0.4 (1.8) |  |  |
|  | Int. week 1 | 0.3 (0.8) | 1.1 (2.8) | 0.8 (0.3 to 1.4) | 0.001 |
| Total physical activity, min | Baseline week | 51.9 (32.7) | 61.8 (47.6) |  |  |
|  | Int. week 1 | 46.0 (32.2) | 63.4 (34.6) | 8.0 (-2.6 to 18.3) | 0.13 |
| Steps | Baseline week | 6982 (2538) | 7831 (3086) |  |  |
|  | Int. week 1 | 6645 (2741) | 8146 (2716) | 741 (55 to 1427) | 0.04 |

Data are presented as means of observed values with standard deviation (SDs) at baseline week and first intervention week in each group. Results from linear mixed model for 140 participants are presented as estimated mean differences (Est. effects) in the intervention group compared with the control group, with corresponding 95% confidence intervals (CIs) and *P*-values. All participants who had at least one observation were included in the analysis. All participants who had at least one observation were included in the analysis. The number of participants with missing data for each variable is 0 to 11 (among *n* = 83) in the control group and 0 to 15 (among *n* = 83) in the intervention group.

### Continuous glucose monitoring

After 7 weeks, there were no statistically significant between-group differences in any glycaemic outcome measures (mean glucose, AUC, CV, and MAGE) derived from CGM across 24-hour, daytime, or nocturnal periods (Table 2). The intervention group showed slightly lower nocturnal glucose AUC than the control group in both the first intervention week (-0.2; 95% CI-0.5 to 0.0, *P =* 0.08) and week 7 (-0.2; 95% CI -0.5 to 0.0, *P =* 0.13), but these differences were not statistically significant (Figure 4).

**Figure 4.**
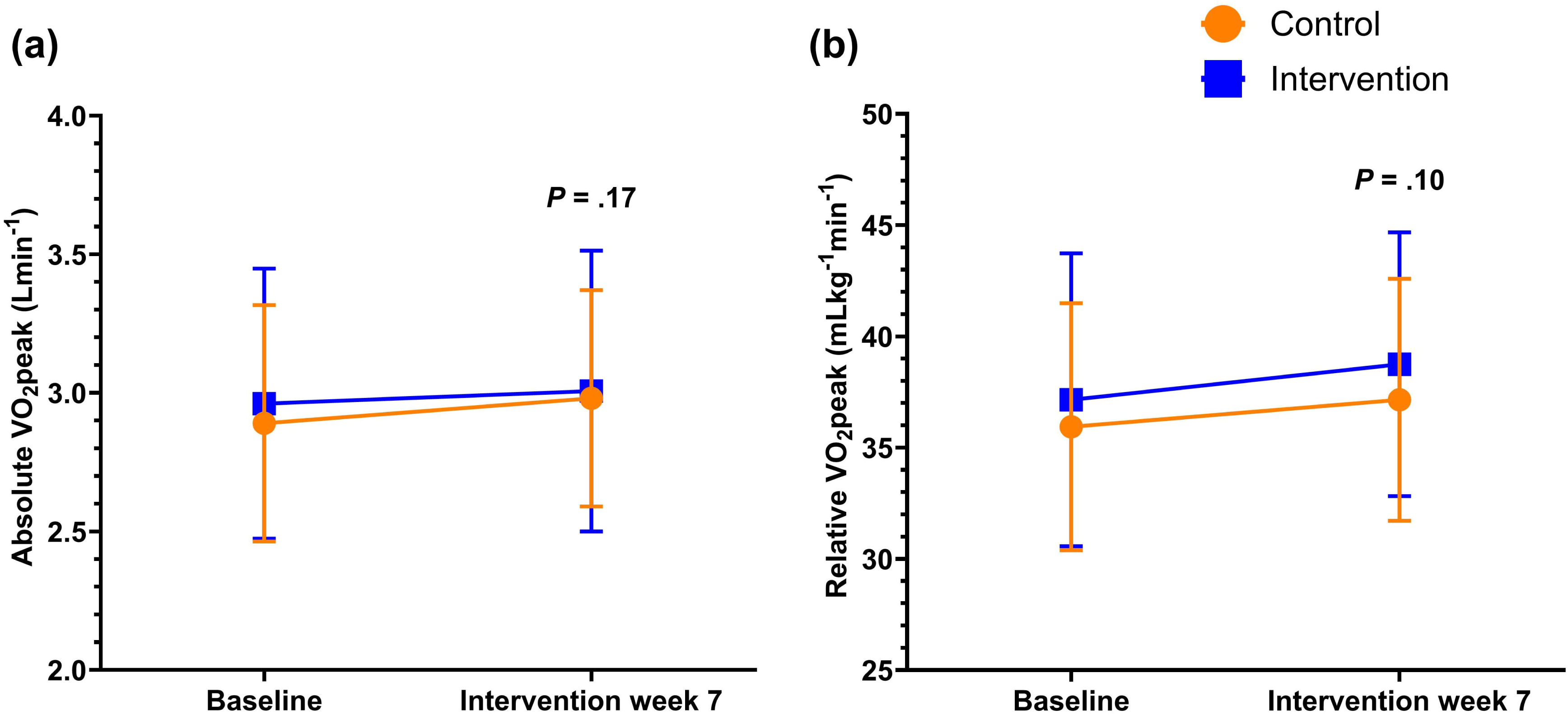
Glucose Area under the curve (AUC) according to group. (a) 24-h glucose AUC, (b) daytime glucose AUC, (c) nocturnal glucose AUC. Data are presented as observed means for intention-to-treat population, error bars show mean ± 1 standard deviation (SD), and the lines represent changes in each group from baseline. The *P*-values were calculated for a test of between group differences, using linear mixed model.

**Table 2.** Glycaemic outcomes estimated from continuous glucose monitoring (CGM) values.

| Outcome | Time | Control | Intervention | Difference<br>(intervention-control) |  |
| --- | --- | --- | --- | --- | --- |
|  |  | Mean (SD) | Mean (SD) | Est. effect (95% CI) | *P* |
| 24-h mean glucose, mmol/L | Baseline week | 4.9 (0.6) | 4.8 (0.5) |  |  |
|  | Int. week 1 | 4.9 (0.5) | 4.9 (0.5) | 0.0 (-0.2 to 0.1) | 0.67 |
|  | Int. weeks 7-8 | 5.0 (0.5) | 5.0 (0.5) | 0.0 (-0.1 to 0.2) | 0.77 |
| Daytime mean glucose, mmol/L | Baseline week | 5.1 (0.6) | 5.0 (0.5) |  |  |
|  | Int. week 1 | 5.1 (0.5) | 5.1 (0.5) | 0.0 (-0.2 to 0.2) | 0.98 |
|  | Int. weeks 7-8 | 5.2 (0.5) | 5.1 (0.5) | 0.0 (-0.1 to 0.2) | 0.61 |
| Nocturnal mean glucose, mmol/L | Baseline week | 4.4 (0.6) | 4.3 (0.5) |  |  |
|  | Int. week 1 | 4.6 (0.5) | 4.4 (0.5) | -0.1 (-0.3 to 0.0) | 0.10 |
|  | Int. weeks 7-8 | 4.6 (0.6) | 4.5 (0.5) | -0.1 (-0.2 to 0.1) | 0.50 |
| 24-h glucose AUC, mmol-h/L | Baseline week | 4.5 (0.5) | 4.4 (0.5) |  |  |
|  | Int. week 1 | 4.5 (0.5) | 4.4 (0.5) | -0.1 (-0.2 to 0.1) | 0.36 |
|  | Int. weeks 7-8 | 4.5 (0.5) | 4.4 (0.5) | 0.0 (-0.1 to 0.2) | 0.76 |
| Daytime glucose AUC, mmol-h/L | Baseline week | 4.9 (0.6) | 4.8 (0.6) |  |  |
|  | Int. week 1 | 4.8 (0.6) | 4.7 (0.5) | 0.0 (-0.2 to 0.1) | 0.73 |
|  | Int. weeks 7-8 | 4.7 (0.6) | 4.7 (0.6) | 0.1 (-0.1 to 0.3) | 0.33 |
| Nocturnal glucose AUC, mmol-h/L | Baseline week | 4.6 (0.7) | 4.6 (0.8) |  |  |
|  | Int. week 1 | 4.7 (0.8) | 4.5 (0.8) | -0.2 (-0.5 to 0.0) | 0.08 |
|  | Int. weeks 7-8 | 4.5 (0.7) | 4.2 (0.8) | -0.2 (-0.5 to 0.1) | 0.13 |
| 24-h CV, % | Baseline week | 16.9 (3.8) | 16.9 (3.8) |  |  |
|  | Int. week 1 | 15.8 (4.0) | 15.9 (3.6) | 0.2 (-1.2 to 1.5) | 0.82 |
|  | Int. weeks 7-8 | 16.6 (5.4) | 17.2 (3.5) | 0.1 (-1.3 to 1.5) | 0.89 |
| Daytime CV, % | Baseline week | 15.7 (3.5) | 15.7 (3.3) |  |  |
|  | Int. week 1 | 15.5 (3.9) | 15.2 (3.6) | -0.4 (-1.7 to 0.8) | 0.51 |
|  | Int. weeks 7-8 | 15.9 (5.3) | 16.4 (3.4) | 0.0 (-1.4 to 1.4) | 0.10 |
| Nocturnal CV, % | Baseline week | 14.1 (5.1) | 14.0 (4.9) |  |  |
|  | Int. week 1 | 11.5 (5.2) | 10.9 (4.3) | -0.3 (-2.1 to 1.5) | 0.74 |
|  | Int. weeks 7-8 | 13.0 (6.9) | 13.8 (5.2) | 0.2 (-1.8 to 2.2) | 0.84 |
| 24-h MAGE, mmol/L | Baseline week | 1.6 (0.4) | 1.6 (0.4) |  |  |
|  | Int. week 1 | 1.6 (0.4) | 1.5 (0.3) | 0.0 (-0.2 to 0.1) | 0.54 |
|  | Int. weeks 7-8 | 1.6 (0.5) | 1.6 (0.3) | 0.0 (-0.1 to 0.1) | 0.96 |
| Daytime MAGE, mmol/L | Baseline week | 1.6 (0.4) | 1.6 (0.4) |  |  |
|  | Int. week 1 | 1.6 (0.4) | 1.5 (0.3) | -0.1 (-0.2 to 0.1) | 0.39 |
|  | Int. weeks 7-8 | 1.7 (0.6) | 1.6 (0.4) | 0.0 (-0.2 to 0.1) | 0.75 |
| Nocturnal MAGE, mmol/L | Baseline week | 1.2 (0.5) | 1.2 (0.4) |  |  |
|  | Int. week 1 | 1.0 (0.4) | 0.9 (0.4) | -0.1 (-0.2 to 0.1) | 0.48 |
|  | Int. weeks 7-8 | 1.2 (0.7) | 1.2 (0.5) | 0.0 (-0.2 to 0.2) | 0.75 |

Data are presented as means of observed values with standard deviations (SDs) at baseline week, intervention week 1, and intervention weeks 7-8. Results from linear mixed model for 150 participants are presented as estimated mean differences (Est. effects) in the intervention group compared with the control group, with corresponding 95% confidence intervals (CIs) and *P*-values. All participants who had at least one observation were included in the analysis. The number of participants with missing data for each variable is 0 to 6 (among *n* = 83) in the control group and 0 to 11 (among *n* = 83) in the intervention group. AUC = Area under the curve, CV = Coefficient of variation, and MAGE = Mean amplitude of glucose excursions.

### Hunger and appetite

After 7 weeks, the intervention participants reported significantly greater hunger, lower satiety, and higher appetite in the evening compared with the controls, with no between-group differences in the morning (Table 4).

**Table 4.**
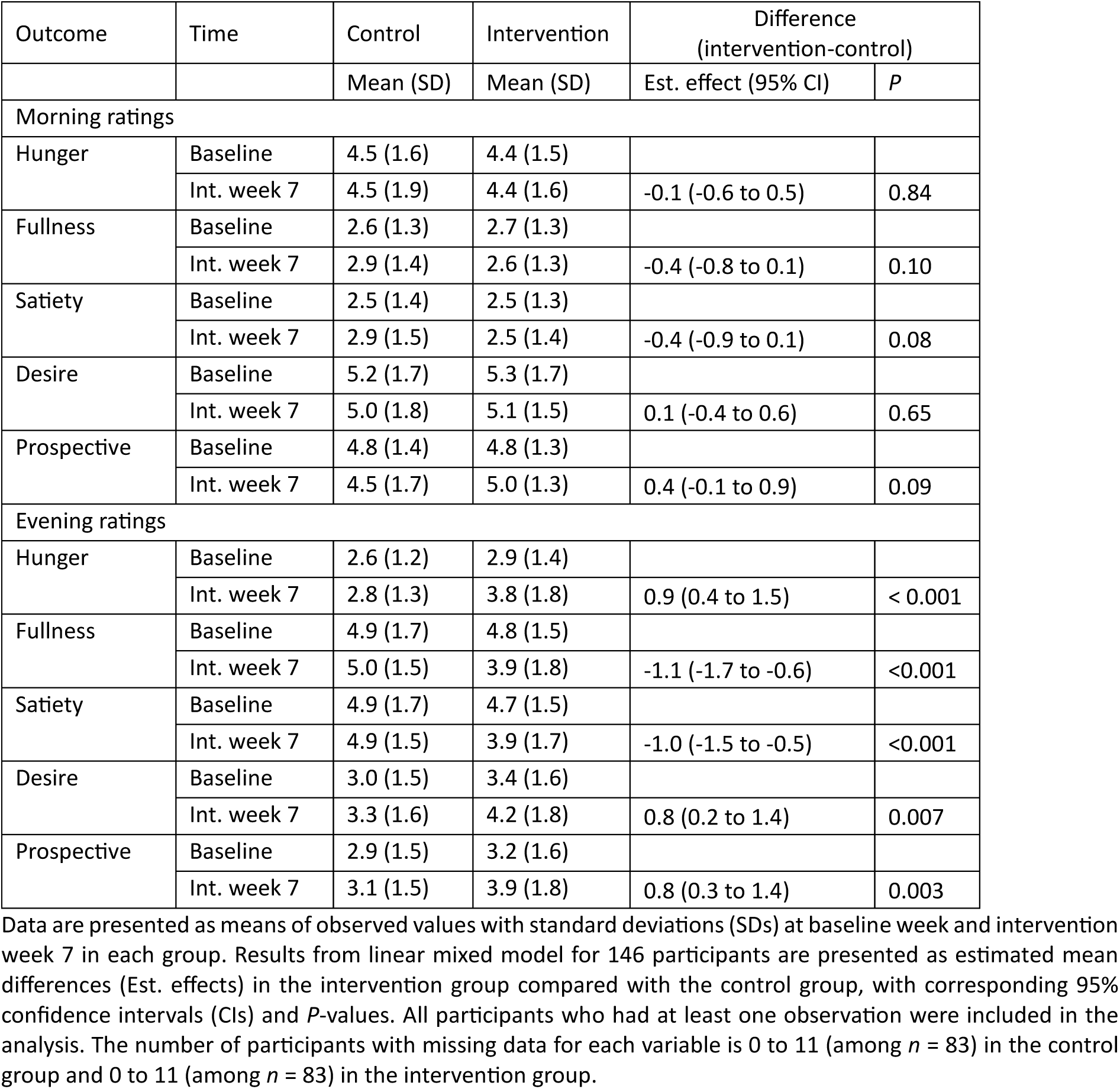
Self-reported ratings of hunger and appetite on visual analogue scale (1-10) in the morning and in the evening in study handbooks.

### Per-protocol analyses

In the per-protocol analyses, we included 39 out of 83 intervention participants (37%) who achieved ≥75 weekly PAI points and reported a ≤10-hour eating window on at least two of four days during the first 7 weeks of preconception intervention. Results from the per-protocol analyses were consistent with those from the intention-to-treat analyses (Supplementary Tables 2-5).

### Adverse events

As reported in the primary article, no serious adverse events occurred during the trial [19]. There were some minor events, including local skin irritation from smartwatches, CGMs, and physical activity monitors. These were resolved by repositioning the devices or, in the case of smartwatches, by replacing plastic with metal armbands.

## DISCUSSION

In this secondary analysis of the BEFORE THE BEGINNING trial, we hypothesised that 7 weeks of preconception exercise combined with TRE would induce greater improvements in cardiorespiratory fitness and glycaemic outcomes compared with a no-intervention control group. Although participants in the intervention group maintained an average daily eating window of ≤ 10 h and met physical activity targets during the preconception period [19], we detected no statistically significant changes in cardiorespiratory fitness or glycaemic outcomes.

In contrast to previous studies, we did not observe improvements in cardiorespiratory fitness parameters (VO₂peak, HR recovery) after 7 weeks. VO₂peak and HR recovery are well-established indicators of cardiorespiratory fitness and higher values in individuals with high BMI is associated with improved insulin sensitivity, reduced visceral and hepatic fat, lower systemic inflammation, and overall better metabolic health [27,28]. Unlike Haganes et al. [15], who combined TRE with three weekly supervised HIIT sessions and observed improvements in VO_2_peak, our intervention was primarily unsupervised, with only two optional supervised sessions. Evidence indicates that supervised exercise promotes adherence and enhances improvements in cardiorespiratory fitness and cardiometabolic outcomes [29]. Our pragmatic, home-based design, intended to reflect real-word conditions, allowed participants to select exercise modality and intensity, supporting long-term feasibility but introducing heterogeneity that may have reduced intervention effectiveness.

Combined TRE and exercise training did not significantly affect CGM-derived glycaemic outcomes. Although not statistically significant, estimated nocturnal glucose AUC was slightly lower in the intervention group after both 1 and 7 weeks, possibly reflecting the effects of prolonged overnight fasting. This finding aligns with the results from a four-arm RCT by Haganes et al. [15], where an 8-week 10-h TRE regimen modestly reduced nocturnal glucose levels.

Previous studies suggest that TRE and high-intensity exercise have greater effects on glycaemic control in metabolically compromised individuals than in metabolically healthy populations [30,31]. Participants in our trial were normoglycaemic at baseline, and CGM-measured glycaemic variability remained within reference ranges [32]. Moreover, reductions in body weight and fat mass, key mediators of improved glucose metabolism [33], were not targeted outcomes and showed minimal change after 7 weeks [19], limiting potential improvements in glycaemic measures. In previous studies showing improved glycaemic outcomes after short-term combined TRE and structured exercise, these improvements were accompanied by favourable changes in body composition [15,34,35].

The duration of the daily eating window may partly explain the limited effects on glycaemic outcomes. Previous studies report improved glycaemic outcomes with ≤ 10 h TRE [15,36], with even greater benefits observed for narrower eating windows (≤ 6 h) [37,38]. In the BEFORE THE BEGINNING trial, we pragmatically selected ≤ 10 h to maximise adherence both before and during pregnancy, allowing participants up to two “off days” per week. While this flexible approach likely supported feasibility and adherence, intermittent compliance may have limited physiological and psychological adaption to fasting.

Physical activity monitoring data from the first week of intervention indicated a small but significant increase in vigorous activity (∼1 min/day) and a non-significant increase in daily steps (∼700 steps/day) in the intervention group, whereas other activity parameters were similar between groups. While any increase in physical activity is beneficial, and increasing volume and intensity leads to greater cardiometabolic benefits [39], the magnitude may have been insufficient to induce measurable cardiometabolic benefits.

Subjective hunger increased in the evening among intervention participants after 7 weeks, while morning hunger remained unchanged. These findings align with a recent meta-analysis data reporting elevated hunger during TRE among people with high BMI, independent of calorie intake [40]. Extended fasting intervals likely contributed to this effect. The flexible TRE protocol, with “off days” may have limited psychological adaptations, in contrast to studies stricter protocols reporting reduced hunger [15].

Limitations of the present study include early conception among 26% (43/166) of participants, resulting in incomplete exposure and missing data at week 7. The mostly home-based and unsupervised intervention nature introduced heterogeneity in exercise type and intensity. We did not monitor physical activity beyond the first intervention week due to limited availability and manufacturer discontinuation of SenseWear armbands, which prevented longitudinal assessment in both groups.

## CONCLUSION

Seven weeks of combined ≤ 10-h TRE and exercise training did significantly improve cardiorespiratory fitness or glycaemic outcomes during the preconception period in people with increased risk of GDM. The intervention’s home-based and unsupervised design reflect real-world conditions but may have limited effectiveness compared with supervised protocols. Baseline normoglycaemia and minimal changes in body composition further constrained potential improvements. Our findings highlight the challenges of implementing effective preconception lifestyle interventions and underscore the importance of developing feasible, targeted strategies for at-risk populations prior to conception.

### What is already known on this topic

- Short-term time-restricted eating combined with supervised exercise training improves body composition, glycaemic outcomes, and cardiorespiratory fitness.

### What this study adds

- Seven weeks of preconception time-restricted eating with largely unsupervised exercise did not improve cardiorespiratory fitness or glycaemic outcomes in people at increased risk of gestational diabetes mellitus.

### How this study might affect research, practice or policy

- Home-based, unsupervised preconception interventions may have limited cardiometabolic impact; supervised or higher-intensity programmes might be needed for measurable benefits.

## Ethical approval

The study was conducted in accordance with the Declaration of Helsinki and approved by The Regional Committees for Medical and Health Research Ethics in Central Norway (REK 143756) on September 24, 2020.

## Data availability statement

Individual deidentified participant data and statistical codes can be found in the Zenodo data repository (https://doi.org/10.5281/zenodo.17709778).

## Author contributions

TM, KÅS, TF, and SLF conceived and contributed to the study design and data analysis plans. GR, MAJS, and HMSS coordinated the study, performed measurements, monitored participants, and supervised the exercise training, with contributions from HNL and APO. MAJS and HMSS drafted the manuscript. All authors provided feedback and approved the final manuscript.

## Funding and acknowledgements

The trial was funded by the Future Leader Award in Diabetes from the European Foundation for the Study of Diabetes and Novo Nordisk Foundation (NNF19SA058975), the Liaison Committee for education, research, and innovation in Central Norway (2020/39645 and 2021/928), and the Joint Research Committee between St. Olavs Hospital and the Faculty of Medicine and Health Sciences, NTNU (FFU, 2021/51833). The funding organisations had no role in study design, data collection, analysis, and publication of results. The authors are thankful to all the participants for their valuable contributions in our study. NeXt Move core facility at the Norwegian University of Science and Technology (NTNU) provided equipment and lab facilities for cardiorespiratory fitness testing and the Clinical Research Facility at St. Olavs Hospital provided clinical measurements. We used eFORSK, which is developed by Central Norway Regional Health Authority for sending out study invitations.

