## Supplementary Table for "Effects of a preconception lifestyle intervention on cardiorespiratory fitness and glycaemic outcomes in people with increased risk of gestational diabetes: Secondary findings from the BEFORE THE BEGINNING randomised controlled trial"

### Supplementary Tables

**Supplementary Table 1. Cardiorespiratory fitness outcomes (Intention-to-treat analysis).**

| Outcome | Time | Control | Intervention | Difference (intervention-control) | |
|---|---|---|---|---|---|
| | | Mean (SD) | Mean (SD) | Est. effect (95% CI) | *P* |
| $VO_2$peak, $Lmin^{-1}$ | Baseline | 2.9 (0.4) | 3.0 (0.5) | | |
| | Int. week 7 | 3.0 (0.4) | 3.0 (0.5) | 0.1 (0.0 to 0.1) | 0.17 |
| $VO_2$peak, $mLkg^{-1}min^{-1}$ | Baseline | 35.9 (5.6) | 37.1 (6.6) | | |
| | Int. week 7 | 37.1 (5.4) | 38.7 (5.9) | 0.8 (-0.2 to 1.9) | 0.10 |
| Rate of perceived exertion | Baseline | 19.2 (0.7) | 19.3 (0.7) | | |
| | Int. week 7 | 19.1 (0.8) | 19.2 (0.7) | 0.0 (-0.2 to 0.3) | 0.83 |
| Respiratory exchange ratio, $VCO_2/VO_2$ | Baseline | 1.11 (0.05) | 1.09 (0.05) | | |
| | Int. week 7 | 1.09 (0.5) | 1.07 (0.05) | -0.02 (-0.03 to 0.0) | 0.06 |
| Heart rate recovery, beats per min | Baseline | 31.2 (11.8) | 30.1 (9.0) | | |
| | Int. week 7 | 32.9 (9.0) | 33.0 (9.6) | 0.6 (-3.1 to 4.3) | 0.76 |

Data are presented as observed means with standard deviations (SDs) at baseline and intervention week 7 in each group. Results from linear mixed model for 163 participants are presented as estimated mean differences (Est. effects) in the intervention group compared with the control group, with corresponding 95% confidence intervals (CIs) and *P*-values. All participants who had at least one observation were included in the analysis. The number of participants with missing data for each variable is 0 to 2 (among *n* = 83) in the control group and 0 to 7 (among *n* = 83) in the intervention group.

**Supplementary Table 2. Cardiorespiratory fitness outcomes (Per-protocol analysis).**

| Outcome | Time | Control | Intervention | Difference (intervention-control) | |
|---|---|---|---|---|---|
| | | Mean (SD) | Mean (SD) | Est. effect (95% CI) | *P* |
| $VO_2$peak, $Lmin^{-1}$ | Baseline | 2.9 (0.4) | 3.0 (0.4) | | |
| | Int. week 7 | 3.0 (0.4) | 3.1 (0.5) | 0.1 (0.0 to 0.2) | 0.07 |
| $VO_2$peak, $mLkg^{-1}min^{-1}$ | Baseline | 35.9 (5.6) | 37.7 (5.2) | | |
| | Int. week 7 | 37.1 (5.4) | 39.4 (4.8) | 1.1 (0.0 to 2.3) | 0.06 |
| Rate of perceived Exertion | Baseline | 19.2 (0.7) | 19.2 (0.7) | | |
| | Int. week 7 | 19.1 (0.8) | 19.1 (0.7) | 0.0 (-0.3 to 0.3) | 0.86 |
| Respiratory exchange ratio, $VCO_2/VO_2$ | Baseline | 1.11 (0.05) | 1.09 (0.05) | | |
| | Int. week 7 | 1.09 (0.5) | 1.06 (0.05) | -0.03 (-0.05 to -0.01) | 0.005 |
| Heart rate recovery, beats per min | Baseline | 31.2 (11.8) | 29.6 (8.8) | | |
| | Int. week 7 | 32.9 (9.0) | 34.8 (8.8) | 2.7 (-1.6 to 6.9) | 0.22 |

Data are presented as observed means with standard deviations (SDs) at baseline and intervention week 7 in each group. Results from linear mixed model for 122 participants are presented as estimated mean differences (Est. effects) in the intervention group compared with the control group, with corresponding 95% confidence intervals (CIs) and *P*-values. All participants who had at least one observation were included in the analysis. The number of participants with missing data for each variable is 0 to 2 (among *n* = 83) in the control group and 0 to 3 (among *n* = 39) in the intervention group.

**Supplementary Table 3. Glycaemic outcomes estimated from continuous glucose monitoring values (Per protocol analysis).**

| Outcome | Time | Control | Intervention | Difference (intervention-control) | |
|---|---|---|---|---|---|
| | | Mean (SD) | Mean (SD) | Est. effect (95% CI) | *P* |
| 24-h mean glucose, mmol/L | Baseline week | 4.9 (0.6) | 4.9 (0.5) | | |
| | Int. week 1 | 4.9 (0.5) | 4.8 (0.5) | -0.1 (-0.3 to 0.1) | 0.17 |
| | Int. weeks 7-8 | 5.0 (0.5) | 5.0 (0.4) | 0.0 (-0.2 to 0.2) | 0.90 |
| Daytime mean glucose, mmol/L | Baseline week | 5.1 (0.6) | 5.0 (0.5) | | |
| | Int. week 1 | 5.1 (0.5) | 5.0 (0.5) | -0.1 (-0.3 to 0.1) | 0.34 |
| | Int. weeks 7-8 | 5.2 (0.5) | 5.2 (0.4) | 0.0 (-0.2 to 0.2) | 0.68 |
| Nocturnal mean glucose, mmol/L | Baseline week | 4.4 (0.6) | 4.4 (0.6) | | |
| | Int. week 1 | 4.6 (0.5) | 4.4 (0.6) | -0.2 (-0.4 to 0.0) | 0.03 |
| | Int. weeks 7-8 | 4.6 (0.6) | 4.4 (0.5) | -0.1 (-0.3 to 0.1) | 0.32 |
| 24-h glucose AUC, mmol·h/L | Baseline week | 4.5 (0.5) | 4.4 (0.5) | | |
| | Int. week 1 | 4.5 (0.5) | 4.4 (0.5) | -0.1 (-0.3 to 0.1) | 0.22 |
| | Int. weeks 7-8 | 4.5 (0.5) | 4.4 (0.5) | 0.0 (-0.2 to 0.2) | 0.91 |
| Daytime glucose AUC, mmol·h/L | Baseline week | 4.9 (0.6) | 4.8 (0.6) | | |
| | Int. week 1 | 4.8 (0.6) | 4.7 (0.5) | -0.1 (-0.3 to 0.2) | 0.59 |
| | Int. weeks 7-8 | 4.7 (0.6) | 4.7 (0.5) | 0.1 (-0.1 to 0.3) | 0.40 |
| Nocturnal glucose AUC, mmol·h/L | Baseline week | 4.6 (0.7) | 4.5 (0.8) | | |
| | Int. week 1 | 4.7 (0.8) | 4.4 (0.7) | -0.3 (-0.5 to 0.0) | 0.07 |
| | Int. weeks 7-8 | 4.5 (0.7) | 4.0 (0.8) | -0.3 (-0.6 to 0.0) | 0.02 |
| 24-h CV, % | Baseline week | 16.9 (3.8) | 16.5 (3.9) | | |
| | Int. week 1 | 15.8 (4.0) | 16.5 (4.3) | 0.8 (-0.8 to 2.5) | 0.32 |
| | Int. weeks 7-8 | 16.6 (5.4) | 17.6 (3.7) | 0.6 (-1.1 to 2.2) | 0.49 |
| Daytime CV, % | Baseline week | 15.7 (3.5) | 15.4 (3.4) | | |
| | Int. week 1 | 15.5 (3.9) | 16.0 (3.9) | 0.4 (-1.1 to 2.0) | 0.60 |
| | Int. weeks 7-8 | 15.9 (5.3) | 16.7 (3.6) | 0.4 (-1.1 to 2.0) | 0.58 |
| Nocturnal CV, % | Baseline week | 14.1 (5.1) | 13.4 (4.7) | | |
| | Int. week 1 | 11.5 (5.2) | 11.9 (4.5) | 0.6 (-1.6 to 2.8) | 0.58 |
| | Int. weeks 7-8 | 13.0 (6.9) | 14.1 (5.2) | 0.7 (-1.5 to 2.9) | 0.54 |
| 24-h MAGE, mmol/L | Baseline week | 1.6 (0.4) | 1.5 (0.4) | | |
| | Int. week 1 | 1.6 (0.4) | 1.5 (0.3) | 0.0 (-0.2 to 0.1) | 0.61 |
| | Int. weeks 7-8 | 1.6 (0.5) | 1.7 (0.3) | 0.1 (-0.1 to 0.1) | 0.55 |
| Daytime MAGE, mmol/L | Baseline week | 1.6 (0.4) | 1.6 (0.4) | | |
| | Int. week 1 | 1.6 (0.4) | 1.6 (0.3) | -0.1 (-0.2 to 0.1) | 0.47 |
| | Int. weeks 7-8 | 1.7 (0.6) | 1.7 (0.4) | 0.0 (-0.2 to 0.2) | 0.84 |
| Nocturnal MAGE, mmol/L | Baseline week | 1.2 (0.5) | 1.2 (0.5) | | |
| | Int. week 1 | 1.0 (0.4) | 1.0 (0.4) | 0.0 (-0.2 to 0.2) | 0.93 |
| | Int. weeks 7-8 | 1.2 (0.7) | 1.3 (0.5) | 0.1 (-0.2 to 0.3) | 0.63 |

Data are presented as means of observed values with standard deviations (SDs) at baseline week, intervention week 1, and intervention weeks 7-8. Results from linear mixed model for 116 participants are presented as estimated mean differences (Est. effects) in the intervention group compared with the control group, with corresponding 95% confidence intervals (CIs) and *P*-values. All participants who had at least one observation were included in the analysis. The number of participants with missing data for each variable is 0 to 6 (among *n* = 83) in the control group and 0 to 2 (among *n* = 39) in the intervention group. AUC = Area under the curve, CV = Coefficient of variation, and MAGE = Mean amplitude of glucose excursions.

**Supplementary Table 4. Daily physical activity outcomes estimated using activity armbands (Per-protocol analysis).**

| Outcome | Time | Control | Intervention | Difference (intervention- control) | |
|---|---|---|---|---|---|
| | | Mean (SD) | Mean (SD) | Est. effect (95% CI) | *P* |
| Physical activity level, METs | Baseline week | 1.3 (0.2) | 1.3 (0.2) | | |
| | Int. week 1 | 1.2 (0.2) | 1.3 (0.1) | 0.0 (-0.1 to 0.07) | 0.67 |
| Daily energy expenditure, kJ | Baseline week | 9347.5 (1332.6) | 9367.9 (1564.8) | | |
| | Int. week 1 | 9369.2 (1293.7) | 9448.1 (1166.1) | 26.9 (-363.1 to 417.0) | 0.89 |
| Sedentary time, min | Baseline week | 1068.1 (157.7) | 1110.8 (117.5) | | |
| | Int. week 1 | 1115.4 (174.8) | 1136.9 (149.4) | 4.9 (-50.1 to 59.9) | 0.86 |
| Light-intensity activity, min | Baseline week | 153.6 (76.7) | 150.7 (61.0) | | |
| | Int. week 1 | 144.8 (77.4) | 144.4 (68.9) | 8.8 (-13.7 to 31.2) | 0.44 |
| Moderate-intensity activity, min | Baseline week | 48.2 (29.4) | 60.0 (44.4) | | |
| | Int. week 1 | 42.3 (29.1) | 58.9 (25.1) | 8.2 (-3.4 to 19.8) | 0.16 |
| Vigorous-intensity activity, min | Baseline week | 3.5 (6.6) | 7.2 (11.4) | | |
| | Int. week 1 | 3.5 (6.7) | 6.1 (5.6) | 0.6 (-2.7 to 3.9) | 0.71 |
| Very-vigorous-intensity activity, min | Baseline week | 0.3 (0.8) | 0.6 (1.7) | | |
| | Int. week 1 | 0.3 (0.8) | 1.2 (3.3) | 0.9 (0.3 to 1.5) | 0.003 |
| Total physical activity, min | Baseline week | 51.9 (32.7) | 67.8 (54.0) | | |
| | Int. week 1 | 46.0 (32.2) | 66.2 (27.4) | 9.6 (-4.4 to 23.5) | 0.13 |
| Steps | Baseline week | 6982 (2538) | 8336 (2754) | | |
| | Int. week 1 | 6645 (2741) | 8323 (2480) | 819 (-13 to 1651) | 0.05 |

Data are presented as means of observed values with standard deviation (SDs) at baseline week and first intervention week in each group. Results from linear mixed model for 102 participants are presented as estimated mean differences (Est. effects) in the intervention group compared with the control group, with corresponding 95% confidence intervals (CIs) and *P*-values. All participants who had at least one observation were included in the analysis. The number of participants with missing data for each variable is 0 to 11 (among *n* = 83) in the control group and 0 to 9 (among *n* = 39) in the intervention group.

**Supplementary Table 5. Self-reported ratings of hunger and appetite on visual analogue scale (1-10) in the morning and in the evening in study handbooks (Per-protocol analysis).**

| Outcome | Time | Control | Intervention | Difference (intervention-control) | |
|---|---|---|---|---|---|
| | | Mean (SD) | Mean (SD) | Est. effect (95% CI) | *P* |
| Morning ratings | | | | | |
| Hunger | Baseline | 4.5 (1.6) | 4.2 (1.5) | | |
| | Int. week 7 | 4.5 (1.9) | 4.5 (1.5) | 0.2 (-0.5 to 0.8) | 0.62 |
| Fullness | Baseline | 2.6 (1.3) | 2.4 (1.2) | | |
| | Int. week 7 | 2.9 (1.4) | 2.5 (1.4) | -0.4 (-0.9 to 0.1) | 0.12 |
| Satiety | Baseline | 2.5 (1.4) | 2.3 (1.3) | | |
| | Int. week 7 | 2.9 (1.5) | 2.5 (1.5) | -0.4 (-0.9 to 0.1) | 0.16 |
| Desire | Baseline | 5.2 (1.7) | 4.7 (1.3) | | |
| | Int. week 7 | 5.0 (1.8) | 5.0 (1.3) | 0.1 (-0.4 to 0.7) | 0.65 |
| Prospective | Baseline | 4.8 (1.4) | 5.0 (1.6) | | |
| | Int. week 7 | 4.5 (1.7) | 5.0 (1.6) | 0.5 (0.3 to 1.1) | 0.04 |
| Evening ratings | | | | | |
| Hunger | Baseline | 2.6 (1.2) | 2.8 (1.3) | | |
| | Int. week 7 | 2.8 (1.3) | 4.2 (1.5) | 1.3 (0.8 to 1.8) | <0.001 |
| Fullness | Baseline | 4.9 (1.7) | 4.6 (1.6) | | |
| | Int. week 7 | 5.0 (1.5) | 3.5 (1.6) | -1.4 (-2.0 to -0.8) | <0.001 |
| Satiety | Baseline | 4.9 (1.7) | 4.4 (1.5) | | |
| | Int. week 7 | 4.9 (1.5) | 3.6 (1.5) | -1.2 (-1.8 to -0.6) | <0.001 |
| Desire | Baseline | 3.0 (1.5) | 3.1 (1.5) | | |
| | Int. week 7 | 3.3 (1.6) | 4.4 (1.7) | 1.1 (0.4 to 1.7) | <0.001 |
| Prospective | Baseline | 2.9 (1.5) | 3.0 (1.4) | | |
| | Int. week 7 | 3.1 (1.5) | 4.3 (1.7) | 1.2 (0.7 to 1.8) | <0.001 |

Data are presented as means of observed values with standard deviations (SDs) at baseline week and intervention week 7 in each group. Results from linear mixed model for 112 participants are presented as estimated mean differences (Est. effects) in the intervention group compared with the control group, with corresponding 95% confidence intervals (CIs) and *P*-values. All participants who had at least one observation were included in the analysis. The number of participants with missing data for each variable is 0 to 11 (among *n* = 83) in the control group and none (among *n* = 39) in the intervention group.
